# A systems immunology analysis of Alzheimer’s disease reveals an age- and environmental exposure-independent disturbance in B cell maturation

**DOI:** 10.1101/2025.06.16.25329692

**Authors:** Stephanie Humblet-Baron, Yang Feng, Rafael Veiga, Julika Neumann, Teresa Prezzemolo, Emanuela Pasciuto, Rik Vandenberghe, Lidia Yshii, Adrian Liston

## Abstract

Alzheimer’s disease is a severe neurodegenerative disorder, with multifactorial mechanisms of disease development and progression. Evidence from genetic association studies, animal models, and clinical investigation suggests a neuroimmunological component to disease, with links to the peripheral immune system. Here we applied a systems immunology approach to determine the immunological correlates of Alzheimer’s disease. Using high-dimensional flow cytometry and machine learning, we comprehensively assessed the cellular component of the peripheral immune system in a well-characterised cohort of 184 Alzheimer’s patients and 105 healthy spouses. Using this approach, Alzheimer’s patients demonstrated a disturbance in B cell maturation, a feature which was associated not only with disease status but also with cognitive decline. This effect was age– and environmental exposure-independent, suggesting a disease-intrinsic relationship between Alzheimer’s disease and B cell maturation. These results provide an under-explored avenue for both improving both mechanistic understanding and therapeutic design in Alzheimer’s disease.

## Introduction

Alzheimer’s disease (AD) is the most common neurodegenerative disorder and a leading cause of dementia, accounting for approximately 60–70% of dementia cases worldwide ^1^. By 2050, the global prevalence of dementia is expected to triple ^2^, underscoring the significant public health challenge posed by AD. Diagnosis of AD is complex, relying on diverse clinical and cognitive criteria, with longitudinal monitoring. The diagnostic challenges of AD are closely related to the complexity of AD pathogenesis ^3^, which involves multiple aspects, including beta-amyloid (Aβ), tau aggregation, neurodegeneration, inflammation, and vascular pathways ^4, 5, 6^. Clinical research in this field has undergone transformative changes over the past years with the *in vivo* biological characterization of Alzheimer disease along the two core axes of amyloid and tau aggregation. The inflammatory and immune-related axis, however, has received comparatively less attention in human studies.

Growing evidence suggests that immune responses are not just secondary to AD pathology but may be a key driving factor to pathogenesis and progression of disease ^7^. The disease biomarkers GFAP and sTREM2, measured in blood or CSF, and TSPO PET, an imaging marker, are primarily associated with innate immune responses ^8, 9, 10^. Genome-wide association studies have revealed that the inflammatory and immune pathway, in both innate and adaptive immunity, are causative in AD. Many AD risk genes associated with immune functions and inflammation have also been identified, including *PSEN2, APOE, CR1, TIM2, TREM2, SPI1, CD33*, and *HLA* ^11, 12, 13, 14, 15^. Variants in these loci are thought to substantially increase the risk of AD via immune activation and diverse immune-related pathways. Beyond genetics, studies using animal models have further illuminated the critical role of immune responses in the trajectory of pathology, with both disease driving and disease inhibition processes. In APP/PS1 mice, knockout of NLRP3 or caspase-1 diminished Aβ plaque formation and increased microglial phagocytosis, protecting from loss of spatial memory and other AD-associated sequelae ^16^. Meanwhile the 5xfAD mouse, when on the Rag-deficient background (deficient in both B and T lymphocytes), showed increased microglial activation, enhanced beta-amyloid pathology and elevated neuroinflammation, demonstrating a protective function of adaptive immunity and suggesting a safeguarding role for antibodies ^17^. These immunological processes are potentially tractable for disease treatment, with several antibody-based therapies targeting amyloid pathology translated into clinical settings in AD, with some demonstrating efficacy and receiving approval for clinical use ^18^. While extensive research underscores the involvement of the immunity in AD, current understanding remains incomplete, in particular regarding the adaptive immune compartment, and requires further research.

Immunophenotyping is a powerful approach of categorising immune cells based on surface/intracellular markers and functional characteristics ^19^. Systems immunology, by integrating high-dimensional data with computational approaches, provides a powerful way to decipher the complex interaction networks within the immune system ^20^. These methods offer promising opportunities to detect early immune cell dysfunctions, facilitating the prediction of diseases, monitoring of progression, and identification of intervention windows well before the appearance of clinical symptoms. Several systems immunology studies have shown associations between dysregulation of homeostasis of peripheral innate and adaptive immunity and markers of cognitive decline ^21^. A recent study reported that increased frequencies of CD8 TEMRA cells and decreased frequencies of naïve CD8 T cells are significantly associated with early β-amyloid accumulation and elevated plasma p-tau181 levels in cognitively-intact individuals ^22^. Another study demonstrated elevation of CD8 TEMRA in the peripheral blood of AD and mild cognitive impairment patients, with increased CD8 TEMRA cells negatively correlated with cognition, while central memory T cells exhibited a positive correlation with cognition ^23^. Park et al., reported the longitudinal increase in B lymphocytes is positively associated with accumulation of cerebral amyloid deposition, and this increase of B cells could also serve as a predictor of Aβ accumulation ^24^. Grayson and colleagues confirmed that several peripheral innate immune cell populations, including mDC and pDC, increased significantly in amyloid-positive mild cognitive impairment participants, while natural killer cells decreased in amyloid-positive cognitively normal participants ^25^.

Despite the potential for systems immunology studies in AD to inform on pathogenesis and improve diagnostics, clinical studies aimed at revealing functional changes in the innate and adaptive immune systems remain limited, particularly with regard to the in-depth analysis of immune cells isolated from peripheral blood ^21^. While large meta-analyses have been performed on basic leukocyte analysis ^26^, few studies have performed high-depth immunophenotyping. The medium-sized cohort studies that have been performed have demonstrated the validity of the peripheral blood as a target for assessment, with changes including lower naïve B cells ^27^ and clonally-expanded CD8 T cells in AD patients ^23^. Here we performed a comprehensive immunological subset analysis, featuring a rich and diverse phenotypic profile of both innate and adaptive immune cells, in a large cohort of 184 AD patients and 105 healthy spouses. High-dimensional flow cytometry and an objective automated gating pipeline was used to leverage immunological data into predictive models for AD. We found diverse immunological changes occurring in AD patients, dominated by the acquisition of a shift in B cell subsets towards the age-associated composition, and loss of coherence in the relationship between B cells and other leukocyte subsets. These results suggest a disease-intrinsic maturational defect in the B cell compartment in Alzheimer’s patients, with potential therapeutic implications.

## Results

### Systems immunology reveals alterations to the peripheral immune system in Alzheimer’s disease patients

To determine the immunological profile associated with the development of AD, we collected peripheral blood mononuclear cells (PBMCs) from a large cohort of well-characterized AD patients (n = 184) and their healthy spousal controls (n = 105). Individuals were characterised for comorbidities and treatments, in addition to AD associated clinical parameters (**Table 1**). PBMCs from each individual were assessed using high dimensional flow cytometry panels, covering extensive T cell and B cell subsets, common myeloid populations, and cytokine production responses to stimulation. Using biologically-relevant pre-defined subsets, 261 subpopulations were quantified for a systems immunology analysis.

**Table 1.** Study population demographics and clinical features.

|  | **Alzheimer's** | **Spouses** |
| --- | --- | --- |
| **n** | 184 | 105 |
| **Age, years<sup>‡</sup> ^** | 70.5±11.3(48-83) | 69.0±13.0 (44-82) |
| **Sex<sup>^</sup>** |  |  |
| Male | 88 | 50 |
| Female | 96 | 55 |
| **Months since diagnosis<sup>‡</sup>** | 24.0±36.0 (0.0-168) | NA |
| **Age at diagnosis (years) <sup>‡</sup>** | 68.0±12.0 (48-79) | NA |
| **APOE-ε4 carriers, n** | 102 | 22 |
| **MMSE** |  |  |
| 25-30 | 33 | NA |
| 20-24 | 48 | NA |
| 10-19 | 80 | NA |
| ≤10 | 23 | NA |
| **Comorbidity, n** |  |  |
| Diabetes | 15 | 11 |
| Autoimmune | 4 | 7 |
| Oncological (inactive) | 11 | 10 |
| Pulmonary | 4 | 4 |
| Other chronic infection | 0 | 5 |
| **Medication, n** |  |  |
| Cholinesterase Inhibitors | 166 | 0 |
| Memantine | 28 | 0 |
| SSRI | 54 | 6 |
| Antipsychotic | 12 | 3 |
| Antiepileptic | 8 | 1 |
| Benzodiazepine | 14 | 5 |
| Cardiovascular | 130 | 51 |
| Anti-aggregants | 57 | 31 |
| Statin | 68 | 34 |
| Osteoporosis | 7 | 0 |
| Stomach Protection | 15 | 5 |
| Thyroid | 14 | 10 |
| Anti-diabetic | 14 | 10 |
| Pulmonary | 14 | 1 |
| Urinary | 10 | 3 |

<sup>‡</sup> Median ± IQR (min-max). MMSE, Mini-Mental State Examination; SSRI, selective serotonin reuptake inhibitors. \* P value, chi-squared test of sex in relation to disease group compared to spouses. ^ non-significant Mann-Whitney U test for age in relation to disease group, compared to spouses.

A multivariate logistic regression identified strong statistical association of multiple PBMC subsets with AD status (**Figure 1A**). Changes in subsets of B cells, CD4 and CD8 T cells, monocytes, NK cells and NKT cells were all associated with disease status, indicating a multifaceted change to the immunological landscape of AD patients. Using a cross-validation approach to identify the parameters with the highest explanatory capacity to discriminate between AD and healthy individuals, the inclusion of top four parameters enhanced explanatory capacity (**Figure 1B**), with an AUC-ROC value of 0.71 (**Figure 1C**). Lower ranked immunological parameters, while individually significant, added no additional information to the multivariate model (**Figure 1B**). Notably, at a global level the composition of the immune system in AD patients was within the normal variation observed in healthy individuals (**Figure 1D**), and it was only when analysis was restricted to the identified high information parameters that immunological divergence between AD and healthy individuals was identified (**Figure 1E**). These results demonstrate that while an immunological signature of AD can be identified from the peripheral blood, it is a discrete dysregulation rather than a global dysfunction.

**Figure 1.**
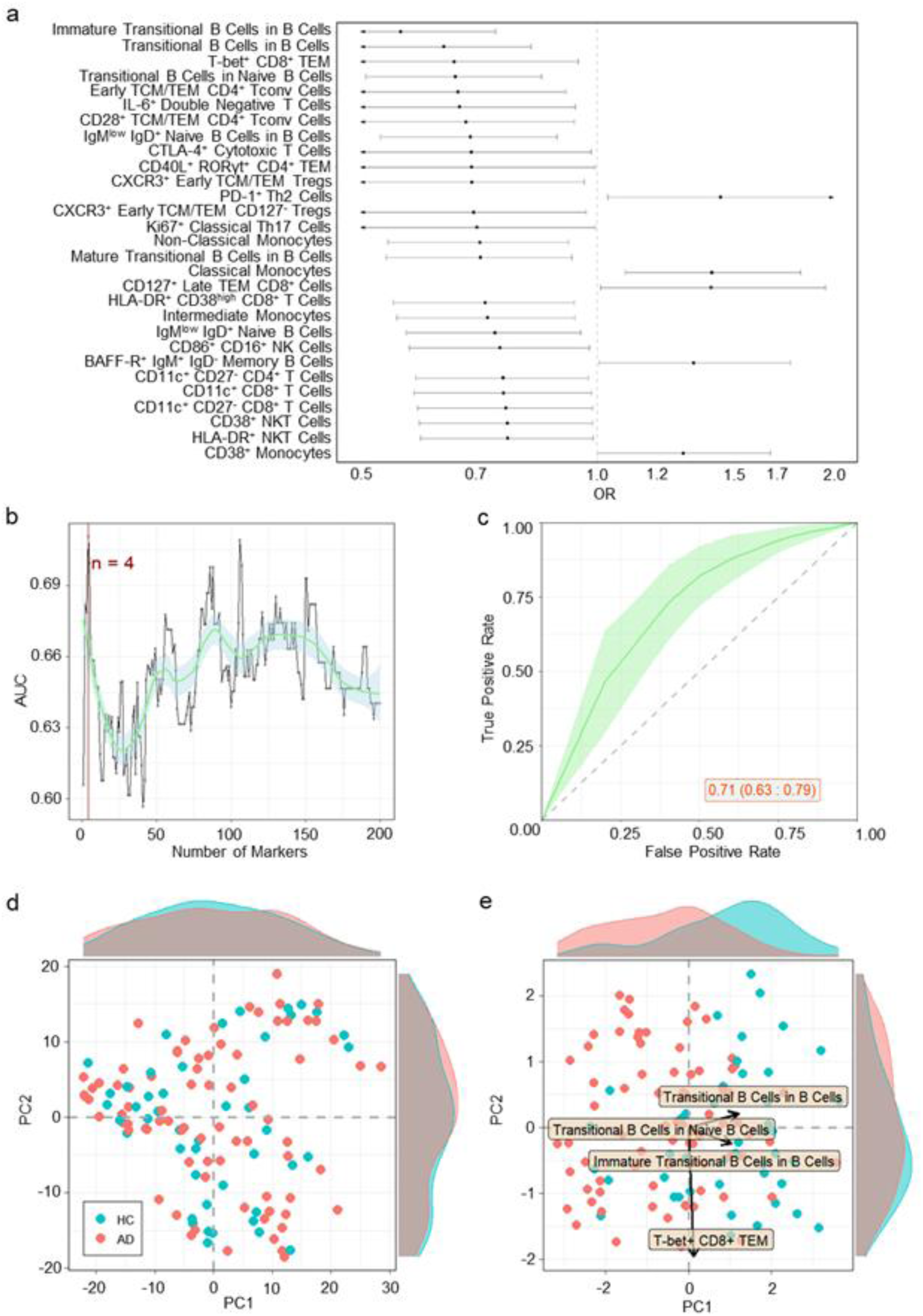
Immunological parameters associated with Alzheimer’s disease. Patients with AD (n=184) were compared to healthy individuals (n=105) by multivariate logistic regression of immunological parameters. **A)** Odds ratio and 95% confidence interval of highly-associated cell population frequency changes in patients with AD in relation to healthy individuals. Estimated by multivariable logistic regression adjusted by sex and age. **B)** Average of 10 fold cross-validation to evaluate a sufficient number of best cell populations, based on ability to adequately discern between AD patients and healthy individuals. **C)** Average ROC curve with 95% confidence interval of 10 fold cross-validation for AD patients in relation to healthy individuals. ROC calculated using multi-variable logistic regression, adjusted by sex and age, considering the 4 cell populations with highest explanatory contribution. Area under ROC curve and confidence interval indicated on graph. **D)** First two PCA components of all cell populations in the dataset. Each dot represents an individual and each colour represents a condition. Histograms show distribution of values in AD patients and healthy individuals. **E)** First two PCA components of 4 cell populations most highly associated for divergence between AD patients and healthy individuals. Histograms show distribution of values in AD patients and healthy individuals. The two arrows show the direction of distinct highly associated cell populations.

### Alzheimer’s disease patients exhibit defective maturation within the B cell compartment

The main drivers of the distinct immunological signature of AD are changes to the B cell compartment. In particular, of the four immunological parameters with the highest explanatory capacity in multivariate models, three are parameters that measure the maturation of the B cell compartment (**Figure 1A**). AD patients have a lower fraction of immature B cells within the naïve B cell compartment and within the total B cell compartment, and in particular have a decrease in the immature (CD10^high^CD21^low^) transitional B cell (CD24^high^CD38^high^) population (**Figure 2**). These interdependent variables are suggestive of a suboptimal developmental pipeline leading from the haematopoietic stem cell compartment in the bone-marrow to the fully mature naïve B cell population found in the circulatory population. The final immunological subset within the top 4 explanatory parameters was the T-bet+ CD8 effector memory population, which was decreased in AD patients compared to healthy controls (**Figure 2**). Among the other associated parameters were changes such as a decrease in CD28+ central memory / effector memory CD4 T cells, increase in PD-1+ Th2 cells, and a slight decrease in non-classical monocytes (**Figure 2**).

**Figure 2.**
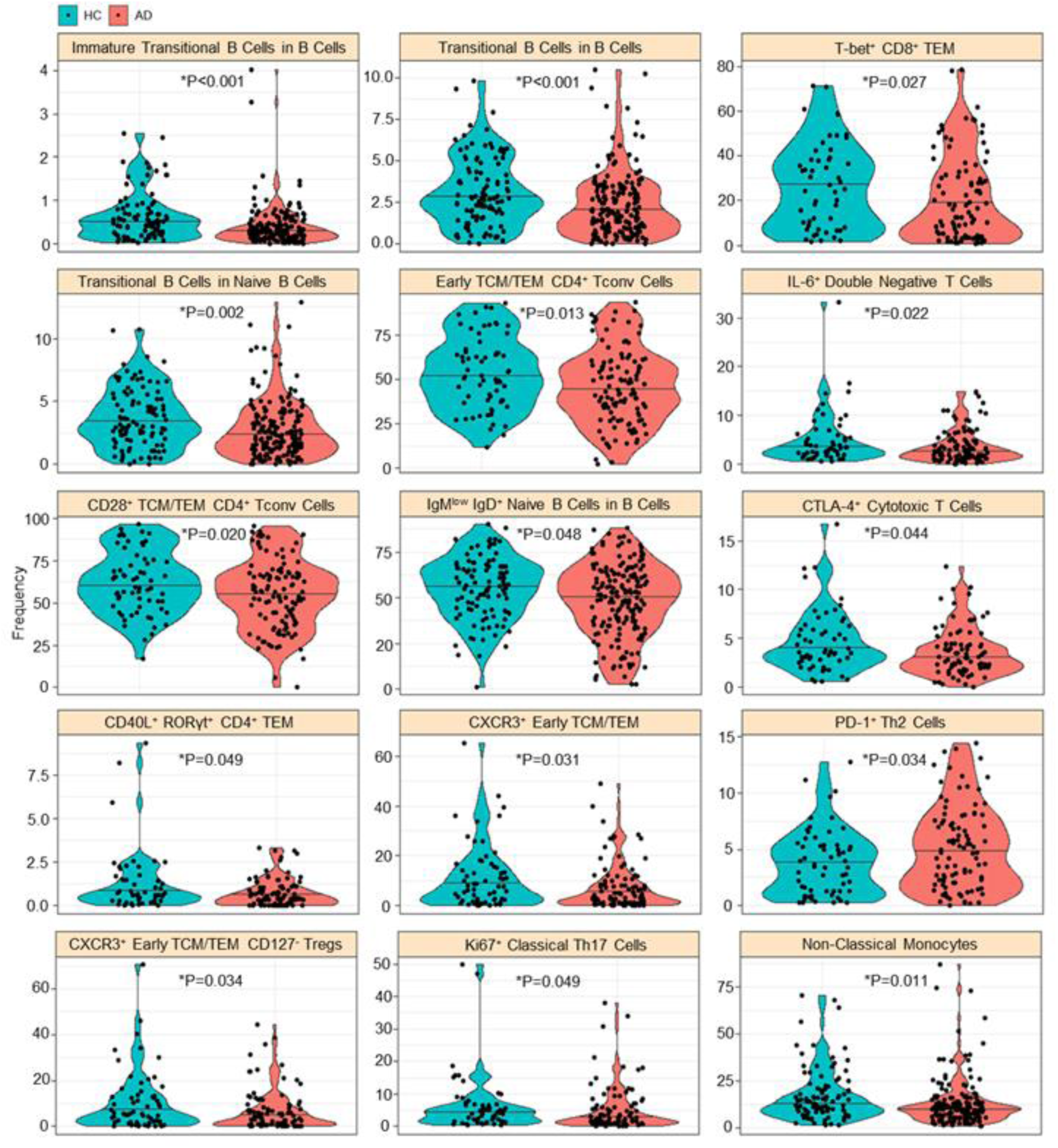
Reductions in naïve and precursor B cell populations in Alzheimer’s disease. Frequency for highly-associated cell populations for AD patients (n=184) and healthy individuals (HC) (n=105). Each dot represents a patient and each colour represents a condition. Raw data values are shown for immature transitional B cells within B cells, transitional B cells within B cells, T-bet+ cells within CD8+ TEM cells, transitional B cells within naïve B cells, early TCM/TEM cells within CD4+ Tconv cells, IL-6+ cells within double negative T cells, CD28+ cells within TCM/TEM CD4+ Tconv cells, IgM^low^IgD+ naïve B cells within B cells, CTLA-4+ cells within cytotoxic T cells, CD40L+ RORγt+ cells within CD4+ TEM, CXCR3+ cells within early TCM/TEM, PD-1+ cells within Th2 cells, CXCR3+ cells within early TCM/TEM CD125-Tregs, Ki67+ cells within classical Th17 cells, and non-classical monocytes within the total monocyte population.

To determine if non-linear relationship between immunological parameters could further boost the discriminatory power of a systems immunology approach, we used a machine learning approach on the immune profiles of AD patients and healthy controls. The strongest discrimination capacity was observed using a Random Forest algorithm. The highest importance parameters (**Figure 3A**) increased discrimination capacity up to a maximum of 0.85 AUC-ROC at 28 parameters (**Figure 3B**), beyond which the information value declined. As with the traditional statistical approach, the machine learning approach identified changes in the maturing B cell compartment as the most important features in discriminating between patients and controls (**Figure 3A**). Lower ranked parameters, which still contributed to the most discriminatory model, were largely T cell subsets, in particular the fraction of CD28+ T cells and proliferating T cell subsets (**Figure 3A**). Compared to the full immunological profile (**Figure 3C**), the 28 parameter phenotype demonstrated stronger divergence between healthy individuals and AD patients (**Figure 3D**).

**Figure 3.**
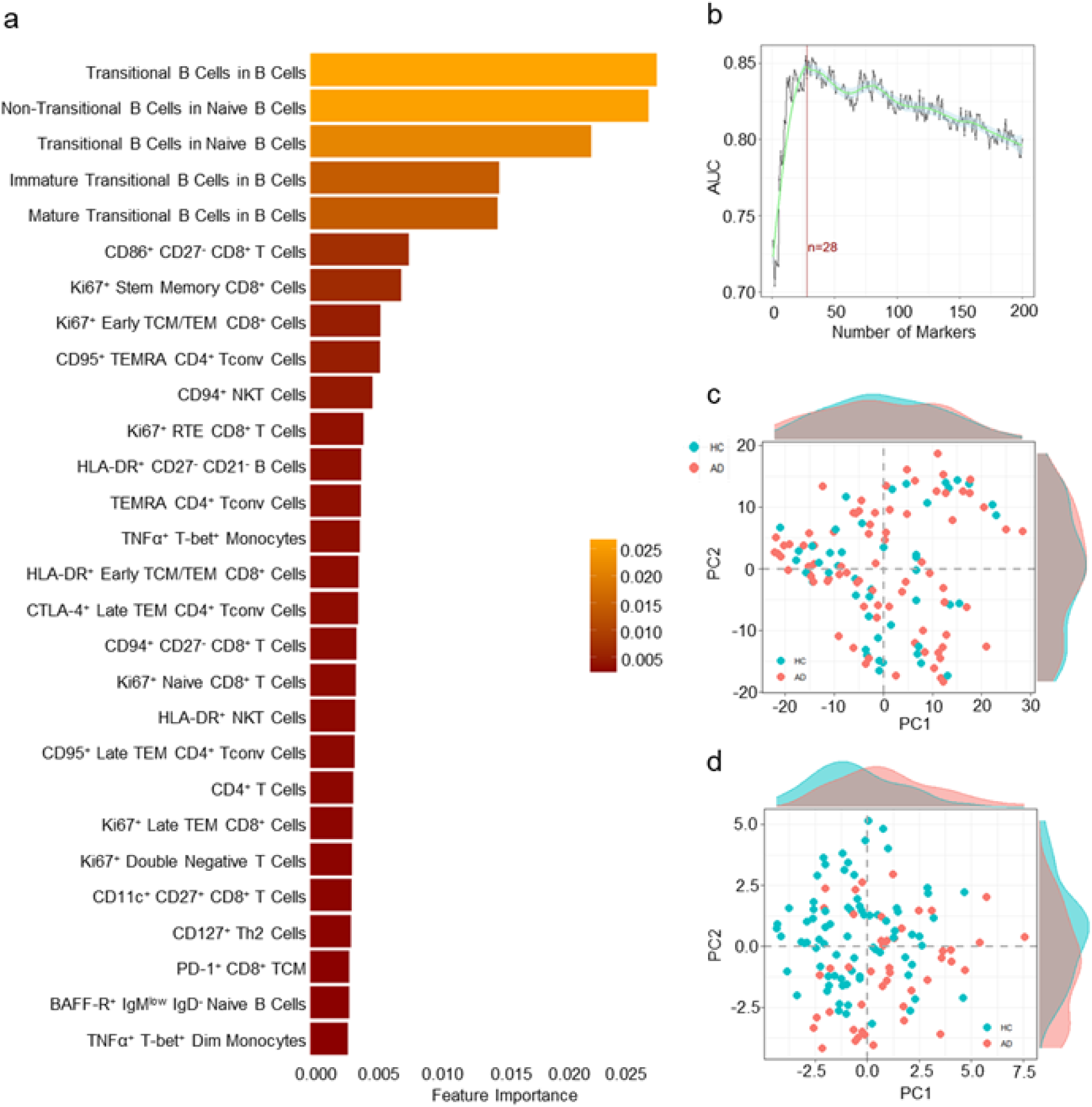
Machine learning-led comparison of Alzheimer’s disease and healthy individuals. A machine learning comparison was performed using immune phenotypes from patients with AD (n=78) and healthy individuals (n=46). **A)** Key immunological features driving machine learning-led disease identification. Model importance for the 28 highest associated cell populations for disease discrimination **B)** Average and 95% confidence interval of 10-fold cross-validation to evaluate enough best cell populations, based on ability to adequately discern between HC and AD. **C)** First two PCA components of all cell populations in the dataset. Each dot represents an individual and each colour represents a condition. Histograms show distribution of values in HC and AD. **D)** First two PCA components of top 28 cell populations most highly associated based on Random Forest for divergence between HC and AD. Histograms show distribution of values in HC and AD.

For further investigation, we considered interactions between the associated immune variables with each other and with clinical characteristics. Considering the inter-relationship of these disease-associated immune parameters, we assessed the correlation between each pair of parameters in healthy individuals, and deviations from these correlations in AD patients (**Supplementary Figure 1**). While the relationship of the B cell maturation parameters with each other were maintained in AD, this relationship was mildly dissociated from many other immune parameters (**Supplementary Figure 1**), indicating dissociation of B cell maturation from the broader immune profile. When AD patients were broken down by APOE ε4 carrier status, no genotype-dependent relationships were observed (**Supplementary Figure 2B**). However association of each immune parameter with MMSE scores, found a significant association with the three B cell maturation parameters and cognitive decline (**Supplementary Figure 2B**). Together these results suggest that the decoupling of B cell maturation capacity from the broader immunological profile is a progressive event in AD, associated with worsening clinical outcomes.

### B cell maturation defects in Alzheimer’s disease are independent of age and environmental exposures

As bone-marrow output of transitional B cells decline with age ^28^, we first tested the hypothesis that defects in the B cell maturational process represented a premature ageing of the immune system. The age structure of our patient and control cohorts was closely matched (average 70.5 years in AD, 69 years in healthy), due to our use of spousal controls (**Figure 4A**), discounting the explanation that immune phenotyping was capturing a demographic difference in the cohort. We further investigated the effect of age within our cohort range (48-83 years in AD, 44-82 years in healthy), calculating the β coefficient of age with each of the most highly associated immunological parameters. Few statistically-significant parameters were associated with age, among either AD patients or healthy individuals (**Figure 4B**). This age-independence was also observed through plotting the immune parameter status, for each of the top variables, with the age of the donor, with the AD-dependent change being observed across the age range (**Figure 4C**).

**Figure 4.**
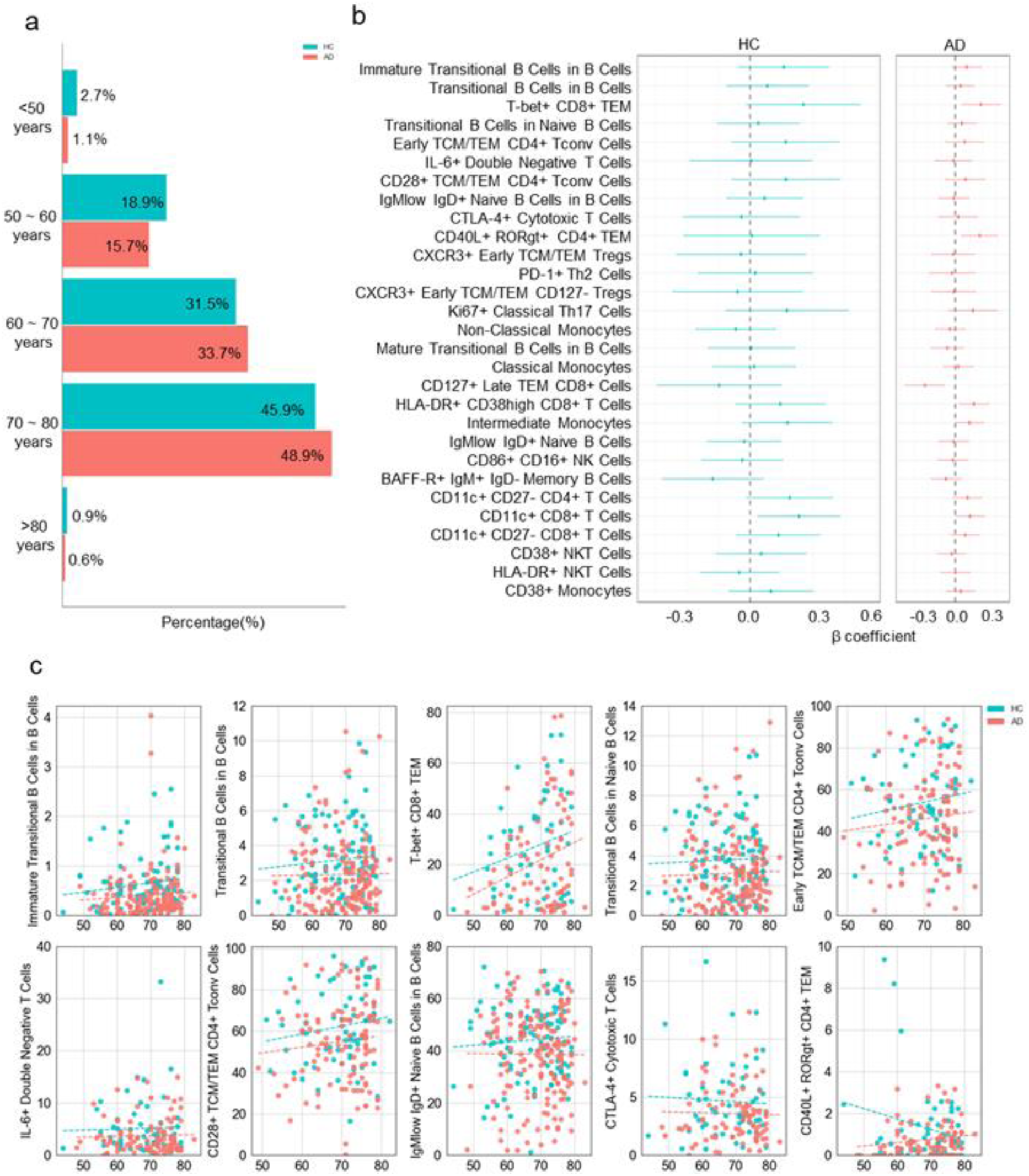
The immunological correlates of Alzheimer’s disease are not driven by age. **A)** Histogram of age structure of cohort, including AD patients (n=184) and healthy controls (n=105). **B)** Forest plots show β co-efficients for age, for the cell populations identified from logistic regression as having an association with AD. β co-efficients were calculated independently for healthy controls and AD patients. **C)** Relationship between the 10 most highly-associated cell populations and age. The dashed line represents the linear regression fit, calculated independently for healthy controls and AD patients.

We next investigated the hypothesis that the altered immune phenotype in AD patients arose from environmental exposures. We started with the seroconversion status against EBV and CMV, based on proposed links to herpesviruses and AD ^29, 30^. In our cohort, no difference in seroprevalence was observed between AD patients and healthy controls (**Figure 5A**). Further, if we limited the analysis to just EBV-seropositive individuals (**Figure 5B**) or CMV-seropositive individuals (**Figure 5C**), all AD-associated immune phenotype changes were maintained, and no significant interaction effects were observed. This suggested the changes observed were not due to differential exposure history to these herpesviruses. To capture the broader impact of environmental exposures, we used the spousal immune phenotype as a reference point. As previously demonstrated ^31^, spouses have a reduced divergence in immune phenotype due to the shared environmental exposures of cohabitation (**Figure 6A**). When comparing spouses to simulated pairings (maintaining the age and sex structure of the pairings), a stronger correlation between spousal values is observed, across a range of immune parameters (**Figure 6B**).

**Figure 5.**
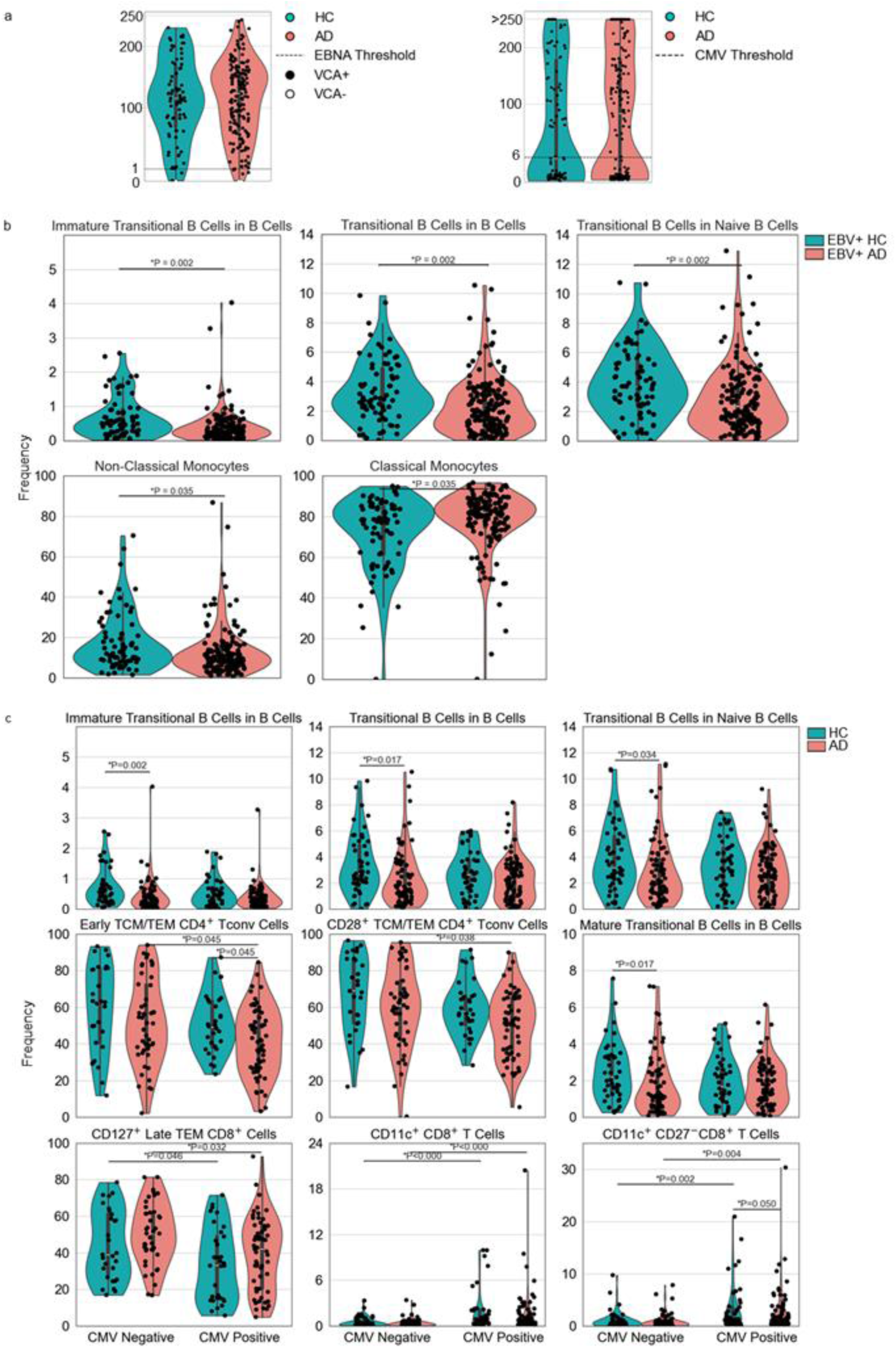
Immunological changes in Alzheimer’s disease patients are independent on EBV and CMV status. **A)** Seropositivity for EBV EBNA-IgG and VCA-IgM (left), and CMV-IgG (right), as determined by ELISA for AD patients (n=184) and healthy controls (n=105). Dashed lines indicate threshold for positivity (EBNA: cut-off=1; CMV: cut-off=6). **B)** EBV seropositive AD and healthy control patients intergroup comparisons were conducted by the Mann-Whitney U test, with the False Discovery Rate (FDR) controlled via the Benjamini-Hochberg method to adjust for multiple comparisons. **C)** Participants were divided into four groups based on CMV status (negative/positive) and disease condition. Kruskal-Wallis test was first applied to detect global differences, followed by pairwise Mann-Whitney U tests with Benjamini-Hochberg correction for multiple comparisons.

**Figure 6.**
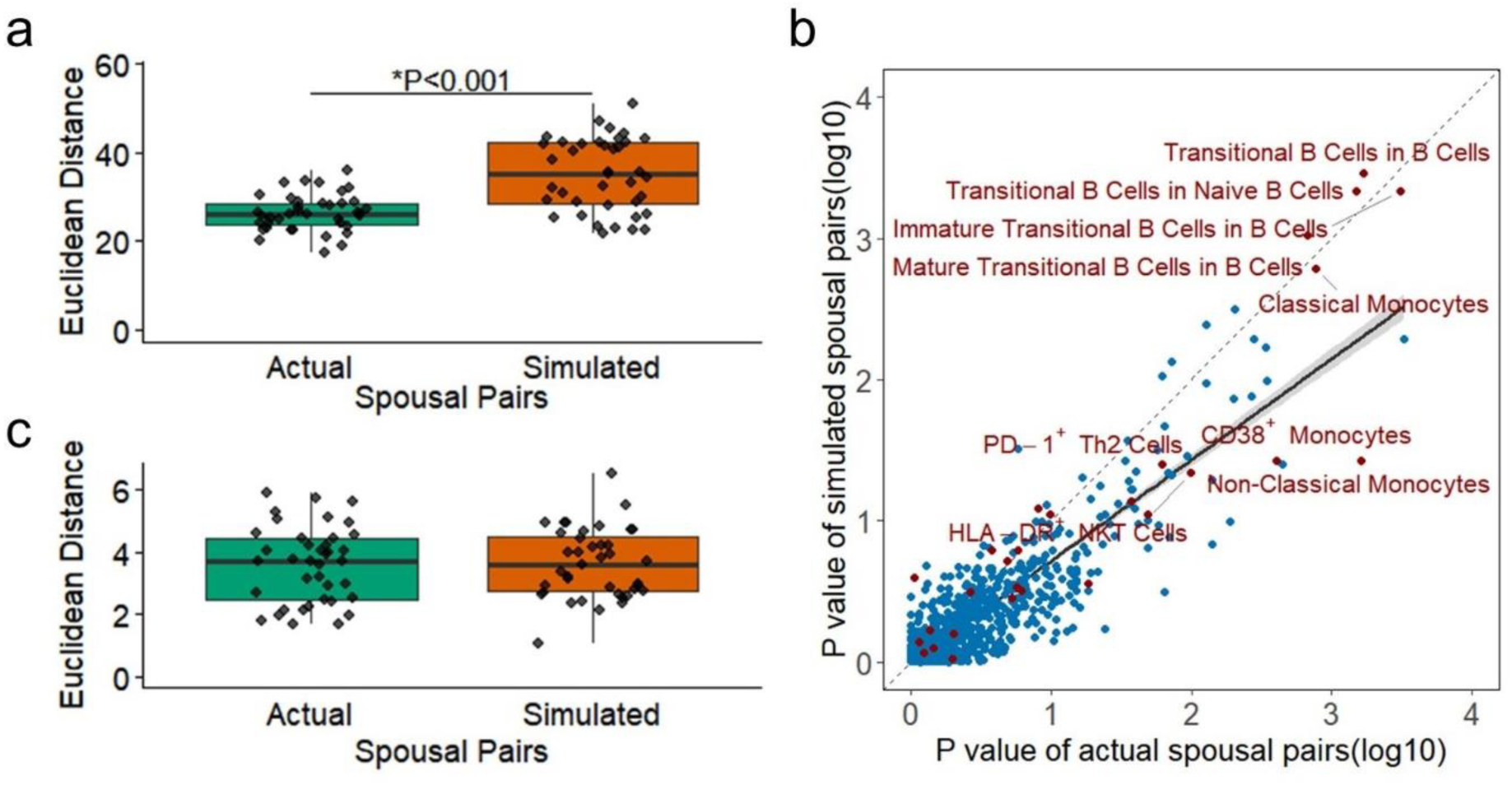
The immunological correlates of Alzheimer’s disease are independent of environmental exposures shared with spouses. **A)** Euclidean distance between spousal pairs, across all immunological parameters. Data calculated for actual spousal pairs, and simulated spousal pairs, randomised within the constraints of maintained sex and age structure (n=39). **B)** Scatter plot of paired p values for the association of each measured immunological parameter. P values were calculated as paired between actual and simulated spousal pairs. Red dots represent the highly-associated parameters of AD. The dashed line indicates the null hypothesis regression, with no effect of shared spousal environment, while the solid line indicates the observed regression. **C)** Euclidean distance between spousal pairs, across only the AD-associated immunological parameters. Data calculated for actual and simulated spousal pairs, as above.

Notably, however, the AD-associated immune phenotypes were largely clustered along the null-relationship line, with equivalent associations observed when simulated pairings were used (**Figure 6B**). Indeed, limiting the immune phenotypes tested to only those associated with AD removed the entire spousal concordance effect (**Figure 6C**). Together, these results suggest that the observed immune changes in AD patients are intrinsic to disease development, rather than emerging from environmental exposures.

## Discussion

The role of the peripheral immune system in the pathophysiology of Alzheimer’s disease is a topic of growing interest, with several mid-sized systems immunology studies identifying links between the peripheral and central immune compartments, including the migration of peripheral CD8+ TEMRA T cells into the CNS ^23, 32^. To further investigate the relationship between the peripheral immune system and AD pathophysiology, we applied a systems immunology approach to a large, well-controlled and well-characterised AD cohort. Our study provides novel insight by identifying specific features of B cell maturation as the most robust immunological changes potentially associated with AD status, as well as multiple immunological changes.

These B cell maturation features showed the strongest association with both disease status, using statistical modelling and machine learning approaches, and with disease severity, as measured by cognitive decline. Our cohort design features, including the use of spousal controls to determine the sensitivity of each feature to environmental exposures, allowed the exclusion of key confounding effects for this association, suggesting that the effect is disease-intrinsic.

In-depth investigation of B cells in AD patients has not been systematically examined. A large meta-analysis of leukocyte counts in the peripheral blood of AD patients reported lower B cell count ^26^, a feature which, while not directly corresponding to our study, identifies the B cell compartment to be perturbed across the broad patient pool. A mid-scale analysis of the B cell compartment identified lower naïve B cells ^27^, a finding which here we have refined to a defect in the CD24^high^CD38^high^ transitional B cell population that serves as an intermediate between the differentiating B cell lineage in the bone marrow and the mature naïve compartment in the peripheral blood. The origin of the lower transitional B cell frequency in AD could derive from several distinct mechanistic processes. The immunosenescence observed in AD patients ^33^, could potentially be reflected in reduced bone marrow output, an enhancement of the diminished bone marrow capacity observed in elderly individuals ^28^. Alternatively, the production may be intact, but the population may be dying off at an abnormal rate. Transitional B cells have recently been reported to be highly susceptible to apoptosis ^34^, and in the neuroinflammatory context of AD ^35^ this vulnerability may be further exacerbated.

Regardless of the origin of the deficit in transitional B cells, this perturbation in B cell maturation may have pathological consequences in AD. A reduction in transitional B cell-mediated turnover of the B cell pool may reduce the efficiency of clonal replacement of existing pathological B cells. A detrimental effect of B cells has been observed in different mouse models, where B cell depletion decreased Aβ plaques and restored cognitive defect ^36^. This effect may in part be mediated through priming and reactivation of neuroinflammatory T cells ^37^. The pathogenic B cell role in mice may be replicated in patients, with Park et al. reporting that a longitudinal increase in total peripheral B cells in Alzheimer’s disease patients over one year correlated with a higher burden of Aβ plaque deposition ^24^. These pathogenic B cell clones may lie within the double negative B cell subset (CD19+IgD-CD27-cells) that, although not identified as altered at the population level in our study, have been shown to be elevated in AD patients as well as in various autoimmune disorders and infectious challenges, as well as during the normal aging process ^27, 38, 39^. Under this model, the defect in B cell maturation would reduce the natural replacement of such pathogenic B cell clones.

An alternative, and not mutually exclusive, model for a role of B cells in the pathophysiology of AD lies in the potential beneficial role of B cells. While the evidence suggests that particular B cell clones may be detrimental, other B cell populations are likely protective during AD. In mice, the constitutive absence of B cells in Rag2-deficient 5xFAD mice drives increased Aβ deposition ^17^, a protective role of B cells that is distinct from the observed punctuated depletion effect ^36^. This may be mediated through the role of rare B cell clones capable of making auto-antibodies targeting Aβ ^40^, a concept translated into clinical practice through ongoing trials with lecanemab (monoclonal anti-Aβ antibody) and related agents ^18^. More likely in this case is the regulatory role of certain B cell subsets, with the production of anti-inflammatory cytokines such as IL-10 and IL-35 protective in AD mouse models ^41, 42^. Interestingly, these regulatory B cell phenotype largely overlap with the transitional B cell population markedly reduced in our AD patients. Under this model, a deficit in transitional B cells in AD patients would contribute to disease through the loss of immunosuppressive cytokines and a subsequent enhancement of neuroinflammatory processes ^43^. Potentially both functions, production of regulatory molecules and replacement of pathogenic clones, could be important protective mechanisms of transitional B cells in AD patients. Both imply a therapeutically-accessible intervention point may be available to prevent or slow disease progression.

While we have focused here on the strongest association, that of B cell maturation, our study identified additional immune deviations in AD patients. Myeloid cell subsets, especially monocytes, are of a particular interest in AD, both due to their close developmental and functional relationship with microglia and because they express the TREM2 AD risk factor. Here we observed an increase in the classical monocyte compartment, a pro-inflammatory cell type, that has been observed in other AD cohorts ^44, 45^. While classical monocytes could contribute to the higher inflammation observed in AD, we did not observe an overall strong activated phenotype in these cells, with the exception of increased CD38 expression, supportive of inflammatory functions ^46^. Complicating the immunological phenotype was a relative deficit in conventional dendritic cells of type 1 (cDC1) and a small decline in the CD4+ type 1 T helper lymphocytes (Th1) and CD8+ type 1 cytotoxic T lymphocyte (Tc1). While classically identified as pro-inflammatory cells due to their role in IFNγ production, IFNγ has a complicated role in the CNS which can include protective functions ^47^.

Beyond the potential mechanistic implications of an immunological understanding of AD, the application of systems immunology approaches provides a potential pathway to earlier diagnosis. Earlier AD diagnosis is considering a high clinical priority, enabling earlier access to care, improved ability to plan for changes in patient needs prior to capacity loss, and earlier access to potentially disease-delaying treatments ^3^. One of the short-comings of diagnostic approaches may be the complex and multifactorial nature of AD pathophysiology, with potential multiple mechanistic paths possible in the development of disease ^48^. The addition of immunological biomarkers, such as the altered B cell maturation identified here, has potential to add to a mixed panel of diagnostic features, covering cognitive, behavioural, neuroimaging, metabolic and neurological components, among overs. Such mixed panel diagnostics may have enhanced robustness and broader utility compared to single-pathway diagnostic approaches ^5^.

### Strengths and limitations of the study

There are several strengths of our study compared to previous deep immunophenotyping efforts. To the best of our knowledge, this represents one of the largest and most comprehensive datasets of deep immunophenotyping in the peripheral blood of AD patients to date, in scale and resolution. The flow cytometry panel was designed to comprehensively capture a wide range of innate and adaptive immune cell subsets, including high-dimensional phenotypic subtypes of T cells, B cells, NK cells, monocytes, dendritic cells, with functional and activation states. The cohort design allowed for a unique assessment of the sensitivity of immune features to shared environmental exposures and enabling the exclusion of key confounding effects. In addition, detailed clinical metadata were collected, including age, sex, cognitive assessment, APOE genotype, and CMV/EBV serological profiling, allowing for robust adjustment and further supporting the interpretation that the observed immunological changes are disease-intrinsic rather than environmentally driven.

Key limitations of our study are the cross-sectional design of the cohort and the prior selection of immunological variables to measure. Due to the single-centre cross-sectional design, where patients were sampled only following disease diagnosis, we cannot determine the causative relationship between the altered immune state and disease. Even the observed association between magnitude of immune change and the extent of cognitive decline would be consistent with either direction of causation. The study was also single-centre in design, relying on statistical cross-validation to identify disease-associated changes, and requires external cohort validation prior to evaluation for use in a diagnostic or therapeutic approach. Furthermore, amyloid or tau biomarker results based on CSF, plasma or PET were available and positive in 102 out of 184 AD cases, while the remaining cases were diagnosed based on clinical-neurological evaluation, neuropsychological assessment, MRI and FDG PET. Another limitation is based on the selection of immunological parameters. While high dimensional flow cytometry enables large-scale immune phenotyping, with enhanced sensitivity for identification of populations with known biological relevance, it does require a prior selection of the phenotypes to be measured. It is therefore possible that additional immune deviations are present in AD patients, but were not covered by our panel design. Additionally, the logistical and ethical constraints of the study prevented sampling of the tissue-resident immune population, and thus it is, in particular, likely that additional alterations to the brain-resident immune system remain to be observed in AD patients. Finally, our study suggests that the observed immune changes are intrinsic to AD rather than being dependent on altered exposure to environmental influences.

While the only environmental influences directly tested were EBV and CMV infection, and untested exposures cannot be excluded from a causative role, the use of spouses to measure environmental effects also exclude untested exposures that would be shared across spouses, including chronic and historical effects.

## Methods

### Study design and patient characteristics

The memory clinic cohort included AD patients (n = 184) and healthy spouse controls (n = 105), recruited from the memory clinic at University Hospitals Leuven, Belgium. The inclusion of the spouse control group was intended to mitigate potential confounding variables, including age, sex, and lifestyle factors, which might influence AD detection outcomes. This approach was designed to enhance the robustness and reliability of comparisons between the AD and control cohorts. Blood samples were collected between 2018 and 2020 following standardized procedures at the memory clinic. 102 AD patients included in the cohort were diagnosed based on amyloid or tau biomarkers, including CSF, blood, molecular PET imaging biomarkers, in accordance with the NIA-AA criteria ^5^, with the remainder diagnosed with clinically probable AD, based on clinical neurological evaluation, neuropsychological assessment, MRI and FDG PET. The inclusion of spouse controls required absence of cognitive impairment or other dementia disorders. The lower age limit was set at 45 years, the upper age limit was 80 years. Exclusion criteria included the presence of significant medical, neurological, or psychiatric comorbidities; the use of immunomodulatory drugs; an active oncological history; current or chronic infections (e.g., hepatitis B, hepatitis C, or HIV); and structural brain lesions evident on MRI, such as large-vessel strokes, sequelae or macro haemorrhages, arachnoid cysts, or posttraumatic lesions. Demographics and clinical characterisation of the study population are given in **Table 1**. Written informed consent was applied to all the participants, in accordance with the Declaration of Helsinki. The study was approved by the UZ/KU Leuven Ethics Committee (S60733).

### Immune profiling of PBMCs

PBMCs from healthy controls and Alzheimer’s disease patients were collected in EDTA tubes and isolated from whole blood by density gradient centrifugation with lymphocyte separation medium (MP Biomedicals) according to the manufacturer’s recommendations. Samples were frozen in FBS with 10% DMSO (Sigma) and stored in liquid nitrogen until the completion of cohort collection. Frozen samples were thawed in complete RPMI, washed and stained with a viability marker (fixable viability dye eFluor780) in the presence of an Fc receptor binding inhibitor cocktail (eBioscience). Cell staining for flow cytometry was performed on three aliquots, as follows:

T cell profiling: staining for anti-CD3 (REA613, Miltenyi Biotec), anti-CD28 (CD28.2, BD Biosciences), anti-ICOS (CD278; DX29, BD Biosciences), anti-CD45RA (HI100, eBioscience), anti-CXCR3 (G025H7, BioLegend), anti-PD-1 (EH12.1, BD Biosciences), anti-CD25 (BC96, BioLegend), anti-CXCR5 (RF8B2, BD Biosciences), anti-CCR2 (CD192; K036C2, BioLegend), anti-HLA-DR (L243, BioLegend), anti-CD31 (WM59, BD Biosciences), anti-IL7Ra (CD127; A019D5, BioLegend), anti-CD8 (SK1, BD Biosciences), anti-CD95 (DX2, BD Biosciences), anti-Ki67 (B56, BD Biosciences), anti-CD4 (SK3, BD Biosciences), anti-FoxP3 (206D, BioLegend), anti-CCR7 (CD197; 3D12, eBioscience), anti-CD14 (TuK4, eBioscience), anti-CTLA-4 (CD152; BNI3, BD Biosciences), anti-CCR4 (CD194; L291H4, BioLegend), and anti-RORγt (Q21-559, BD Biosciences).

B cell profiling: staining for anti-CD3 (REA613, Miltenyi Biotec), anti-CD123 (7G3, BD Biosciences), anti-CD80 (L307.4, BD Biosciences), anti-CD57 (HNK-1, BioLegend), anti-CD21 (B-ly4, BD Biosciences), anti-CD27 (L128, BD Biosciences), anti-CD24 (ML5, BioLegend), anti-BAFF-R (11C1, BD Biosciences), anti-CD94 (HP-3D9, BD Biosciences), anti-HLADR (L243, BioLegend), anti-CD19 (HIB19, BioLegend), anti-IgM (MHM-88, BioLegend), anti-CD8 (SK1, BD Biosciences), anti-CD86 (2331 (FUN-1), BD Biosciences), anti-CD141 (1A4, BD Biosciences), anti-CD56 (NCAM16.2, BD Biosciences), anti-CD4 (SK3, BD Biosciences), anti-CD16 (3G8, BD Biosciences), anti-CD40 (5C3, BD Biosciences), anti-CD11c (3.9, BioLegend), anti-IgD (IA6-2, BioLegend), anti-CD14 (TuK4, eBioscience), anti-CD10 (HI10a, BioLegend), and anti-CD38 (HB-7, BioLegend).

Cytokine profiling: To assess intracellular cytokine production, cells were first cultured for 4h in the presence of phorbol mysristate acetate (1 µg/ml, Sigma-Aldrich), ionomycin (1 µg/ml, Sigma-Aldrich), and brefeldinA (BD), followed by staining for anti-CD3 (REA613, Miltenyi Biotec), anti-IL2 (MQ1-17H12, BD Biosciences), anti-TNFα (MAb11, BioLegend), anti-CD40L (CD154; 24-31, BioLegend), anti-PD-1 (EH12.1, BD Biosciences), anti-CD25 (BC96, BioLegend), anti-IL10 (JES3-9D7, BD Biosciences), anti-4-IBB (CD137; 4B4-1, BioLegend), anti-HLA-DR (L243, BioLegend), anti-CD19 (HIB19, BioLegend), anti-IL-4 (MP4-25D2, BioLegend), anti-CD8 (SK1, BD Biosciences), anti-IFNγ (4S.B3, BD Biosciences), anti-Tbet (4B10, BD Biosciences), anti-CD45RA (HI100, BD Biosciences), anti-CD4 (SK3, BD Biosciences), anti-Gata3 (L50-823, BD Biosciences), anti-IL-17a (N49-653, BD Biosciences), anti-FOXP3 (206D, BioLegend), anti-CCR7 (CD197; 3D12, eBioscience), anti-CD14 (TuK4, eBioscience), anti-CTLA-4 (CD152; BNI3, BD Biosciences), anti-IL-6 (MQ2-13A5, BD Biosciences), and anti-RORγt (Q21-559, BD Biosciences).

Cells were fixed using the Foxp3/Transcription factor buffer staining set (eBioscience). All samples were run on a BD FACSymphony A3 Cell Analyzer (BD Biosciences). Data were compensated with AutoSpill ^49^ and manually pre-processed to exclude acellular events. Identification of leukocyte populations and marker expression was performed in R (version 4.0.2) with a custom script (Neumann *et al*., manuscript in preparation), resulting in the populations listed in **Supplementary Spreadsheet 1**.

### Systems immunology analysis

Gating calculated the frequency of 2389 pre-defined leukocyte populations or subpopulations. Variation in low-frequency subsets made the calculation of sub-populations from these subsets difficult, due to missing data. To maintain comparability between individuals and consistency within the database, we excluded cell populations with more than 20% missing data. Consequently, data analysis was performed on a set of 761 immune variables (cell population frequencies).

### Normality Transformation and Standardization

After removing variables with missing data, Box-Cox transformations were applied to each immune variable to improve normality. For each variable, a minimal shift of 1 was subtracted to ensure all values were strictly positive. The optimal Box-Cox lambda parameter was estimated using the BoxCox.lambda() function from the forecast package in R (version 4.3.3), using the log-likelihood method with bounds set between –5 and 5. If lambda equalled 0, a natural logarithm was applied instead. Following Box-Cox transformation, all immune variables were standardized using z-score normalization. For each variable, the mean and standard deviation were calculated across all samples, and values were rescaled to have a mean of zero and a standard deviation of one.

### Data Imputation

The remaining variables were then imputed using k-nearest neighbor (kNN) imputation via the kNN() function from the VIM package in R (version 4.3.3). Prior to imputation, all markers were transformed and normalised to comparable scales and distributions; kNN was then performed using Euclidean distance with k = 3 neighbours.

### Multivariate models

Multivariate models were applied to investigate associations between immune cell population frequencies and clinical or demographic variables. Logistic regression models were constructed for each immune variable, with disease status (AD/control) as the outcome and both age and sex included as covariates. For each model, odds ratios (OR), 95% confidence intervals (CI), and p-values were extracted to quantify the association between the immune variable and disease. Linear regression models were used to examine associations between immune variables and age. Each immune variable was regressed on standardized age and sex. Regression coefficients (β), confidence intervals, and p-values were reported for the age term.

### Penalised multivariate logistic regression

To assess predictive performance and immunological independent join effect, we applied penalised logistic regression with L2 penalization on the imputed dataset. Models were built incrementally by including the top N variables (N = 1…200) according to the univariate ranking, each time adjusting for age and sex. Performance was evaluated by stratified 10-fold cross-validation, computing the mean area under the receiver operating characteristic curve (AUC). We observed that AUC increased up to four variables, beyond which generalisation declined.

### Immune-phenotypic similarity by principal component analysis

Overall immune similarity between individuals was examined via principal component analysis (PCA) on Euclidean distances calculated across all 934 imputed variables. The first two principal components did not clearly separate Alzheimer’s patients from controls. Repeating PCA using only the four most disease-associated markers revealed a distinct clustering of cases and controls. To aid interpretation, vectors corresponding to each of these four markers were projected onto the PCA biplot.

### Machine learning

Random Forest (RF) classifiers were employed to capture potential complex relationships among markers. Maximum tree depth was constrained to improve generalisation. For each fold of a stratified 10-fold cross-validation, the optimal maximum depth was selected based on (OOB) score (analogous to a nested CV procedure). The most frequently chosen depth was then used to train a final model on the full dataset. Feature importance was assessed by the Gini impurity decrease, ranking immune variables by their contribution to classification. To assess how the number of variables influences model generalisation, RF classifiers were then constructed incrementally using the top 1 to 200 ranked immune variables by Gine score. Stratified 10-fold cross-validation was applied to calculate the AUC for each feature count.

### Data and code availability

Raw data for the study is available from https://data.mendeley.com/datasets/Liston cohort AD ST1 Part A (DOI: 10.17632/y7bknr9whz.1); ST1 Part B (DOI: 10.17632/5wp4n6v7y2.1); ST2 Part A (DOI: 10.17632/z4ywrgz4nw.1); ST2 Part B (DOI: 10.17632/5ry7dxpsf4.1); ST2 Part C (DOI: 10.17632/6t7pzgsdbh.1); ST3 Part A (DOI: 10.17632/276kmc6sht.1), ST3 Part B (DOI: 10.17632/8b8fh2b88x.1).

Source data is available as **Supplementary Spreadsheet 2**. Source code is available on GitHub at: https://github.com/rafael-veiga/AlzhemiersDisease.

## Data Availability

Raw data for the study is available from https://data.mendeley.com/datasets/Liston cohort AD ST1 Part A (DOI: 10.17632/y7bknr9whz.1); ST1 Part B (DOI: 10.17632/5wp4n6v7y2.1); ST2 Part A (DOI: 10.17632/z4ywrgz4nw.1); ST2 Part B (DOI: 10.17632/5ry7dxpsf4.1); ST2 Part C (DOI: 10.17632/6t7pzgsdbh.1); ST3 Part A (DOI: 10.17632/276kmc6sht.1), ST3 Part B (DOI: 10.17632/8b8fh2b88x.1).
Source data is available as Supplementary Spreadsheet 2. Source code is available on GitHub at: https://github.com/rafael-veiga/AlzhemiersDisease.

## Acknowledgments

We thank the patients and volunteers who made the study possible. This work has received funding from the European Union’s Horizon 2020 research and innovation program under grant agreement No 874707 (EXIMIOUS), UZ Leuven Klinische Onderzoeks-en Opleidingsraad, VLAIO (Flanders Innovation & Entrepreneurship) with the project entitled PRISMA and by Stichting Alzheimer Onderzoek – Fondation Recherche Maladie Alzheimer (SAO-FMA). The authors acknowledge the important contributions of Carine Schildermans, Jeason Haughton, Mathijs Willemsen and the KU Leuven FACS Core. The authors declare no conflict of interest.

## Supplementary Figures

**Supplementary Figure 1.**
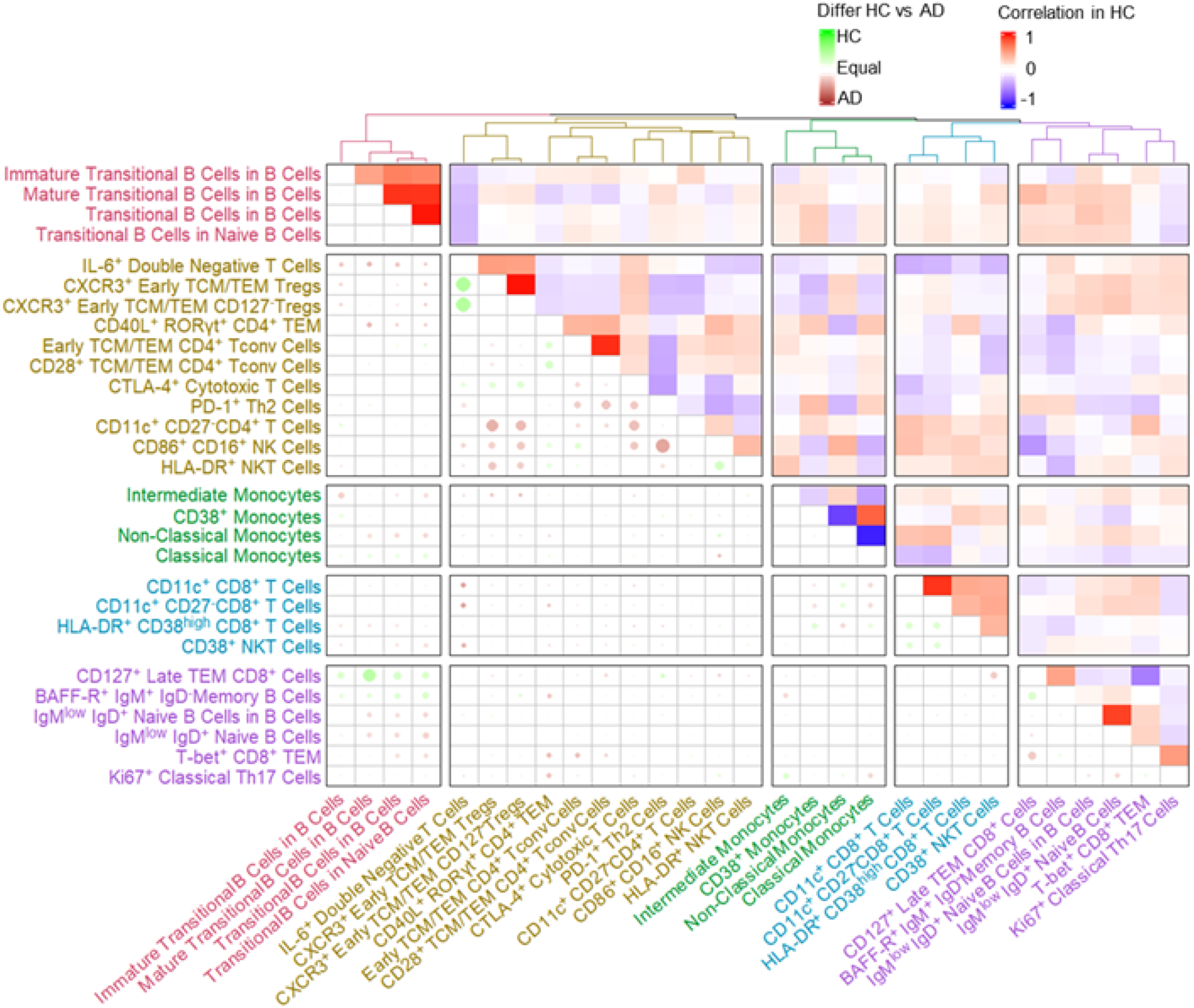
Distortions in the immunological relationships present in Alzheimer’s disease patients. The correlation coefficients were calculated between each pair of AD-associated immunological parameters. Correlations present in the healthy population (top-right) are shown, with colour indicating the strength and direction of the observed correlation. Deviations present in AD patients are shown in the bottom right, with colour and size indicating the strength and direction of correlation changes, from the baseline set in healthy individuals. No value indicates preservation of the relationship observed in healthy individuals.

**Supplementary Figure 2.**
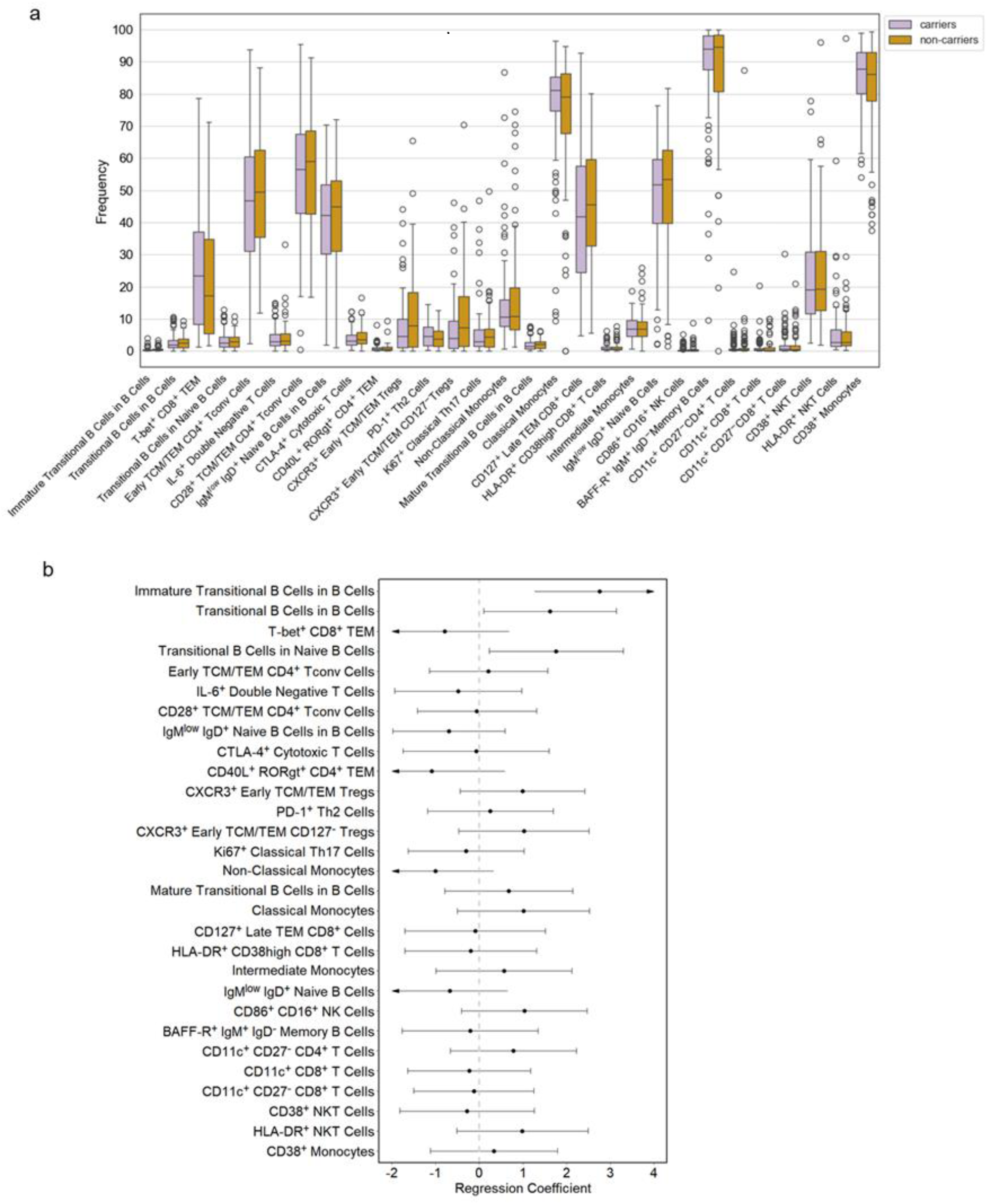
Genotype and disease severity associations of immunological changes in Alzheimer’s disease. **A)** AD patients were segregated based on APOE-ε4 genotype (102 carriers, 82 non-carriers), and assessed for genotype-dependent immune phenotype effects. Group differences were evaluated by a two-sided Mann–Whitney U test and p values were adjusted for multiple comparisons using the Benjamini–Hochberg procedure. **B)** Forest plot of Tobit regression estimating the association between the frequency of highly-associated cell populations and Mini-Mental State Examination (MMSE) scores, adjusted by sex and age.

